# Medication Improves Age Disparities in Six-Month Treatment Retention for Opioid Use Disorder

**DOI:** 10.1101/2020.02.21.20023143

**Authors:** Carrie M. Mintz, Ned J. Presnall, John M. Sahrmann, Jacob T. Borodovsky, Paul E.A. Glaser, Laura J. Bierut, Richard A. Grucza

**Affiliations:** Department of Psychiatry, Washington University School of Medicine, St Louis, MO USA; Department of Internal Medicine at Washington University School of Medicine, St Louis, MO USA

**Keywords:** opioid use disorder, adolescence, buprenorphine, naltrexone, treatment retention, age disparity, insurance disparity

## Abstract

**Background and Aims:** Adolescents with opioid use disorder (OUD) are an understudied and vulnerable population. We examined the association between age and six-month treatment retention, and whether any such association was moderated by medication treatment.

**Methods:** In this retrospective cohort study, we used an insurance database with OUD treatment claims from 2006-2016. We examined 261,356 OUD treatment episodes in three age groups: adolescents (ages 12-17), young adults (18-25) and older adults (26-64). We used logistic regression to estimate prevalence of six-month retention before and after stratification by treatment type (buprenorphine, naltrexone, or psychosocial services only). Insurance differences (commercial vs Medicaid) in medication treatment prevalence were also assessed.

**Results:** Adolescents were far less likely to be retained compared to adults (17.6%; 95% CI 16.5-18.7% for adolescents; 25.1%; 95% CI 24.7-25.4% for young adults; 33.3%; 95% CI 33.0-33.5% for older adults). This disparity was markedly reduced after adjusting for treatment type. For all ages, buprenorphine was more strongly associated with retention than naltrexone or psychosocial services. Adolescents who received buprenorphine were more than four times as likely to be retained in treatment (44.5%, 95% CI 40.6-49.0) compared to those who received psychosocial services (9.7%, 95% CI 8.8-10.8). Persons with commercial insurance were more likely to receive medication than those with Medicaid (73.3% vs 36.4%, χ^2^ =57,870.6, (*p*<.001).

**Conclusions:** Age disparities in six-month treatment retention are strongly related to age disparities in medication treatment. Results point to need for improved implementation of medication treatment for persons with OUD, regardless of age or insurance status.

## 1. Introduction

As the opioid epidemic continues, there has been increased interest in understanding outcomes in adolescents with opioid use disorder (OUD) (Borodovsky et al., 2018; Camenga et al., 2019; Hadland et al., 2018; Hadland et al., 2017). As with other substance use disorders, the risk of OUD often begins in adolescence (Chen et al., 2009; Miech et al., 2015; World Health Organization, 2009), and opioid-related morbidity and mortality has increased within the adolescent population in recent years (Gaither et al., 2016; Gaither et al., 2018; Groenewald et al., 2019), emphasizing the need for effective treatments for this population.

Pharmacologic treatments for OUD include three FDA-approved medications: buprenorphine, methadone and naltrexone, which exists in oral and injectable extended-release forms. Studies in adult populations have shown that treatment with buprenorphine, methadone or extended-release naltrexone is superior to non-pharmacologic treatment for treatment retention (Dole and Nyswander, 1965; Fudala et al., 2003; Lee et al., 2018; Mattick et al., 2003), and that buprenorphine and methadone in particular decrease risk of opioid overdose (Larochelle et al., 2018; Morgan et al., 2019; Sordo et al., 2017).

Increasing data show pharmacologic treatment is also beneficial for youth, often inclusively defined as adolescents and young adults. For example, buprenorphine is associated with treatment retention (Marsch et al., 2005; Marsch et al., 2016; Woody et al., 2008) and decreased illicit opioid use (Marsch et al., 2005; Marsch et al., 2016). In addition, receipt of any OUD medication has been associated with better treatment retention compared to psychosocial services among persons aged 13-22 (Hadland et al., 2018). Recognizing OUD medication effectiveness in youth, the American Academy of Child and Adolescent Psychiatry now offers buprenorphine waiver training during its annual meeting (American Academy of Child and Adolescent Psychiatry Annual Meeting, 2019) and the American Academy of Pediatrics has recommended increased access to medication treatment for adolescents with OUD (Committee On Substance Use and Prevention, 2016).

However, adolescents are less likely than their adult counterparts to receive OUD medication (Hadland et al., 2017): It is estimated that fewer than 5% of adolescents with OUD receive medication treatment (Feder et al., 2017; Hadland et al., 2018). Reasons for this low prevalence are multifactorial. There are well-known barriers to prescribing medication for OUD that are likely enhanced for providers who treat adolescents. For example, federal regulations require methadone be dispensed by certified opioid treatment programs, and there are additional restrictions for those younger than 18 (Wachino and Hyde, 2015). Buprenorphine is FDA-approved for those 16 years of age or older, but physicians must complete eight hours of training in order to obtain the required Drug Enforcement Administration waiver. The waiver training has been previously cited as a barrier by physicians (Huhn and Dunn, 2017) and may be perceived as particularly cumbersome to providers who treat adolescents, for whom OUD is relatively rare. Naltrexone is FDA approved for those 18 years or older in both the oral and extended-release injectable formulations, and may be prescribed in an office setting without additional required training or waivers. However, the oral formulation has not been shown to be efficacious in the adult population (Minozzi et al., 2011) and the extended-release formulation, which was approved for OUD in 2010, while potentially beneficial for treatment retention (Lee et al., 2018), has not been shown to decrease overdose risk (Morgan et al., 2019). In addition, the extended-release formulation can be prohibitively expensive depending on insurance status (Substance Abuse and Mental Health Services Administration, 2014).

Finally, despite growing data demonstrating benefit of medication treatment for youth with OUD, there remains a relative lack of outcomes data in the adolescent population specifically (World Health Organization, 2009). To our knowledge, no study has examined whether OUD medications show similar effectiveness in adolescent populations as they do in the adult population. Further, no study has examined associations between different medications and treatment retention within the adolescent population. Increased knowledge in these areas may help guide treatment decisions with respect to medication utilization for adolescents with OUD.

Administrative claims databases offer several advantages when studying treatment outcomes of adolescents with OUD. Specifically, they allow for large sample sizes that would be impossible to obtain via a randomized control trial. They include data from different geographic regions and in naturalistic settings, improving the generalizability of the results. Finally, they allow for the identification and examination of long-term outcomes. Primary outcomes in OUD treatment studies vary and include both direct (e.g., urine drug testing) and indirect measures of sobriety. A commonly used indirect measure of treatment effectiveness is retention in care (Hadland et al., 2018; Presnall et al., 2019; Williams et al., 2019). Importantly, treatment retention is a clinically important outcome that has been associated with improved sobriety (Kakko et al., 2003; Mattick et al., 2014) and decreased morbidity (Schwartz et al., 2009; Woody et al., 2014) and mortality (Larochelle et al., 2018; Sordo et al., 2017) in persons with OUD.

Thus, the objectives of this study were to 1) examine whether age is associated with six-month OUD treatment retention, 2) whether treatment with OUD medication moderates the relationship between age and retention 3) compare different medication treatments for OUD with respect to treatment retention within the adolescent population. We used a large administrative database composed of commercial and Medicaid insurance holders to examine six-month treatment retention of three age groups: adolescents (12-17 years old), young adults (18-25 years old) and older adults (26-64 years old). First, we examined the association between age and treatment retention. Second, we examined the effect of treatment type on the association between age and six-month retention. Finally, we compared retention rates of each treatment type within each age group. Based on previous studies in adult and youth populations, we hypothesized adolescents with OUD would be less likely to be retained in treatment at six months compared to adults, but that after adjusting for medication treatment, this disparity would decrease. In addition, we predicted that similar to what has been previously observed in adult populations, six-month treatment retention would be more strongly associated with buprenorphine than with naltrexone or non-pharmacologic treatment within the adolescent population.

## 2. Methods

### 2.1 Data Source

Data were from the MarketScan Commercial Claims and Encounters and Medicaid databases (Truven Health Analytics, Ann Arbor, Michigan). The commercial database includes claims for employees, spouses, and their dependents from several large employers and health plans; the Medicaid database includes claims from enrollees from eleven states. States from which Medicaid data were provided were unknown to authors. Both databases provide claims for inpatient and outpatient health care services and prescription drugs. Data were de-identified prior to receipt and were thus deemed exempt from human subjects review by the Institutional Review Board at Washington University School of Medicine. As data were not considered human subjects, the requirement for patient consent was obviated.

### 2.2. Sample Selection

Persons with OUD were identified between January 1, 2006 and December 31, 2016 in the commercial database and between January 1, 2011 and December 31, 2016 in the Medicaid database using International Classification of Diseases (ICD)-9 and ICD-10 codes (Supplementary Table 1). Supplementary Figure 1 details exclusion criteria at the level of the individual, including no prescription drug coverage, treatment claims occurring outside periods of continuous enrollment, and those occurring exclusively in setting of hospitalization(s).

**Table 1.** Demographics of Study Sample.

|  | All Ages |  | 12-17 |  | 18-25 |  | 26-64 |  |
| --- | --- | --- | --- | --- | --- | --- | --- | --- |
|  | <u>Commercial</u> | <u>Medicaid</u> | <u>Commercial</u> | <u>Medicaid</u> | <u>Commercial</u> | <u>Medicaid</u> | <u>Commercial</u> | <u>Medicaid</u> |
|  | *n*=150,900 | *n*=126,184 | *n*=3,219 | *n*=1,792 | *n*=57,441 | *n*=21,026 | *n*=90,240 | *n*=103,366 |
| Sex |  |  |  |  |  |  |  |  |
| Mean (SD) | 33.7 (12.8) | 33.9 (9.7) | 16.3 (1.0) | 16.0 (1.1) | 21.8 (2.0) | 22.7 (2.1) | 42.0 (10.1) | 36.4 (8.8) |
| Male | 93,643 (62.1) | 44,365 (35.2) | 1,933 (60.1) | 1,052 (58.7) | 38,447 (66.9) | 6,022 (28.6) | 53,263 (59.0) | 37,291 (36.1) |
| Female | 57,257 (37.9) | 81,819 (64.8) | 1,286 (40.0) | 740 (41.3) | 18,994 (33.1) | 15,004 (71.4) | 36,977 (41.0) | 66,075 (63.9) |
| Race |  |  |  |  |  |  |  |  |
| White | n/a | 102,405 (83.2) | n/a | 1,332 (75.3) | n/a | 17,982 (87.3) | n/a | 83,091 (82.5) |
| Black | n/a | 8,823 (7.2) | n/a | 215 (12.2) | n/a | 895 (4.3) | n/a | 7,713 (7.7) |
| Hispanic | n/a | 1,414 (1.2) | n/a | 43 (2.4) | n/a | 214 (1.0) | n/a | 1,157 (1.2) |
| Other | n/a | 10,499 (8.5) | n/a | 180 (10.2) | n/a | 1,512 (7.3) | n/a | 8,807 (8.7) |
| Co-morbid SUD | 56,351 (37.3) | 55,898 (44.3) | 1,922 (59.7) | 1,264 (70.5) | 27,020 (47.0) | 9,871 (47.0) | 27,409 (30.4) | 44,763 (43.3) |
| Co-morbid psychiatric disorder | 77,819 (51.6) | 69,852 (55.4) | 2,277 (70.7) | 1,358 (75.8) | 29,189 (50.8) | 10,285 (48.9) | 46,353 (51.4) | 58,209 (56.3) |
| Previous OUD diagnosis | 68,206 (50.4) | 67,147 (53.2) | 1,207 (37.5) | 468 (26.1) | 30,371 (52.9) | 11,058 (52.6) | 36,628 (40.6) | 55,621 (53.8) |
| Treatment Type |  |  |  |  |  |  |  |  |
| Psychosocial only | 40,264 (26.7) | 80,211 (63.6) | 2,206 (68.5) | 1,635 (91.2) | 18,585 (32.4) | 14,421 (68.6) | 19,473 (21.6) | 64,155 (62.1) |
| Buprenorphine | 95,476 (63.3) | 29,916 (23.7) | 689 (21.4) | 65 (3.6) | 33,105 (57.6) | 4,720 (22.5) | 61,682 (68.4) | 25,131 (24.3) |
| Methadone | 2,882 (1.9) | 12,846 (10.2) | 29 (0.9) | 3 (0.2) | 832 (1.5) | 1,406 (6.7) | 2,021 (2.2) | 11,437 (11.1) |
| Naltrexone (oral) | 9,788 (6.5) | 1,821 (1.4) | 277 (8.6) | 86 (4.8) | 3,510 (6.1) | 243 (1.2) | 6,001 (6.7) | 1,492 (1.4) |
| Naltrexone (XR) | 2,490 (1.7) | 1,390 (1.1) | 18 (0.6) | 3 (0.2) | 1,409 (2.5) | 236 (1.1) | 1,063 (1.2) | 1,151 (1.1) |

SUD= substance use disorder. OUD= opioid use disorder. XR= extended release. Data presented as n (percent) except for age, for which mean and SD are shown.
Numbers do not always sum to 100.0 due to rounding.

**Figure 1.**
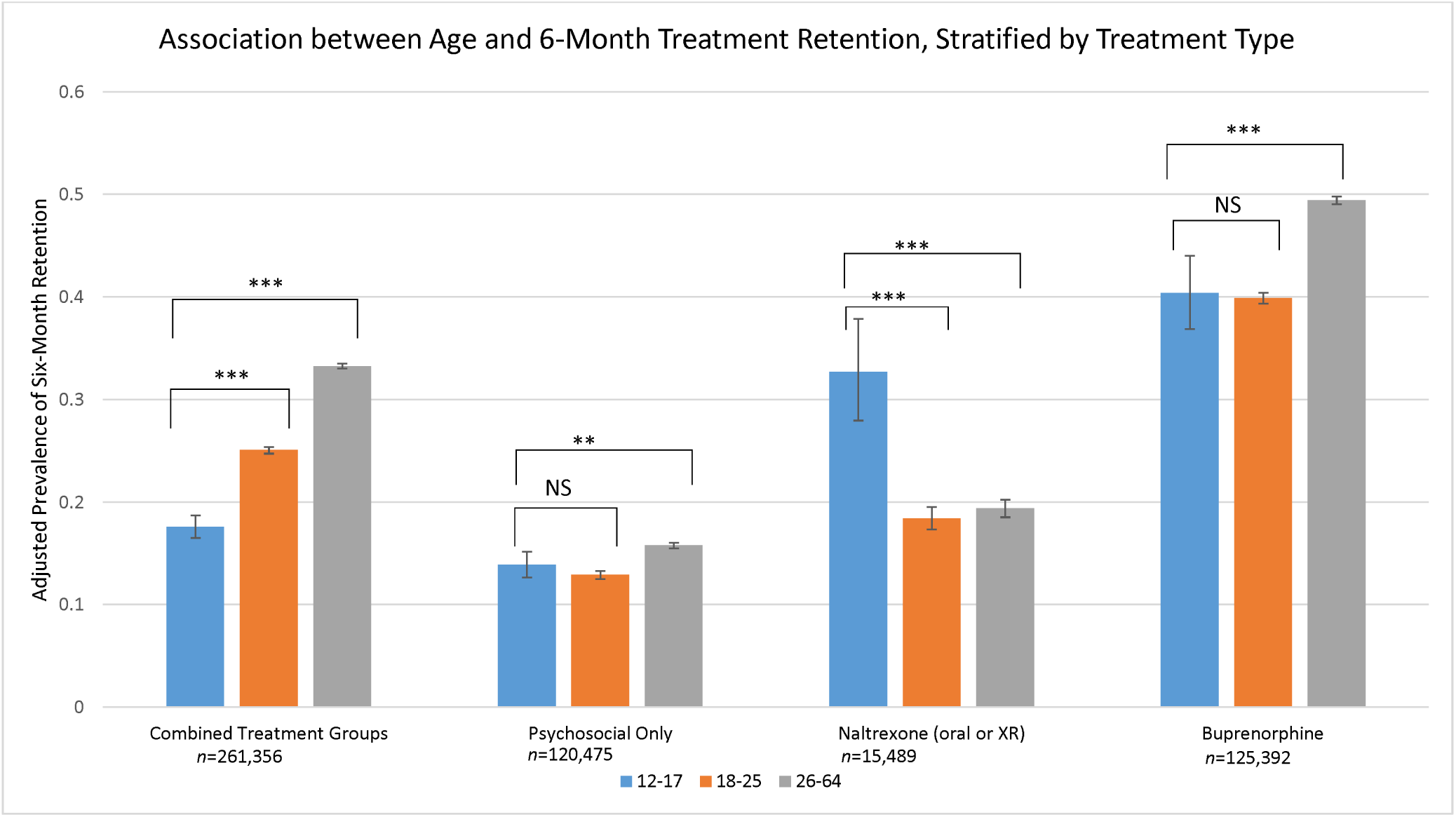
All models adjusted for sex, co-morbid substance use disorder, co-morbid psychiatric disorder, previous diagnosis of opioid use disorder, insurance status and year. XR=extended-release. Error bars represent 95% confidence intervals. NS= Non-significant at the *p*<.05 level. ^**^Comparison significant at the *p*<.01 level; ^***^Comparison significant at the *p*<.001 level.

OUD treatment services were characterized using Current Procedural Terminology codes, the Healthcare Common Procedure Coding System (HCPCS), and ICD-9 and ICD-10 codes (Supplementary Table 2). Buprenorphine treatment—either as a monoproduct or combined with naloxone—was identified using pharmacy claims that included an appropriate National Drug Code (NDC). Treatment with oral naltrexone was identified using pharmacy claims that included appropriate NDCs; extended-release naltrexone treatment was identified with pharmacy claims with relevant NDCs and the HCPCS code J2315. Methadone treatment was identified using HCPCS code H0020. NDCs used to describe pharmacologic treatment are detailed in Supplementary Table 3.

**Table 2.** Predicted prevalence of six-month retention by treatment type, stratified by age group.

|  | <u>12-17</u> | <u>18-25</u> | <u>26-64</u> |
| --- | --- | --- | --- |
|  | *n*=4,979 | *n*=76,229 | *n*=180,148 |
|  | % (95% CI) | % (95% CI) | % (95% CI) |
| Buprenorphine | 44.49 (40.63, 49.03) | 41.51 (40.95, 42.07) | 49.19 (48.79, 49.58) |
| Naltrexone (oral or XR) | 31.73 (27.05, 36.82) | 19.58 (18.49, 20.71) | 21.47 (20.61, 22.35) |
| Psychosocial services only | 9.74 (8.82, 10.75) | 10.74 (10.42, 11.08) | 18.10 (17.82, 18.37) |

All comparisons significant at $p < .001$ . CI= confidence interval. XR= extended release. All models adjusted for sex, co-morbid substance use disorder, co-morbid psychiatric disorder, previous diagnosis of opioid use disorder, insurance status and year.

Our unit of analysis was the treatment episode. We defined a treatment episode as starting with receipt of any OUD treatment (pharmacologic or psychological), and terminating when a gap of at least 45 days in without a treatment claim was observed. We chose a 45 days as this connoted at least two weeks without medication receipt, which was determined to be synonymous with treatment drop out. Each treatment episode was required to be preceded by at least six months of medical and pharmacy coverage to collect data on covariates. In addition, it was required that each treatment episode be associated with medical and pharmacy coverage that lasted at least six months after the start of the treatment episode. Individuals could undergo multiple treatment episodes within the data set. Supplementary Figure 1 details treatment episode exclusion criteria.

### 2.3 Variables of Interest

Primary outcome was treatment retention at six months. Treatment type consisted of the following categories: psychosocial services only, oral naltrexone, extended-release naltrexone, methadone, and buprenorphine. Treatment episodes that included both psychosocial services and medication were counted in the medication group. Treatment episodes that included more than one kind of pharmacotherapy were excluded from analyses. Age was treated as a categorical variable with three levels: 12-17 years old, 18-25 years old, and 26-64 years old.

Covariates included sex, year of treatment episode, OUD diagnosis prior to current treatment episode, insurance status, co-morbid substance use disorder(s) (SUD) and co-morbid non-SUD psychiatric disorder(s). Presence of a co-morbid SUD was measured as a dichotomous variable and included the following use disorders: alcohol, sedative, cocaine, cannabis, amphetamine, and “other.” Non-SUD psychiatric co-morbidity was measured as a dichotomous variable and included the following: depressive disorder, bipolar disorder, psychotic disorder, anxiety disorder, somatoform disorder, eating disorder, conduct disorder, developmental disorder, personality disorder, other mental health disorder, and other child psychiatric disorder. Co-morbidities were identified from inpatient and outpatient claims using the ICD-9 and ICD-10 codes (Supplementary Table 1). Co-morbid SUDs and non-SUD psychiatric disorders were included if there was a relevant diagnosis made in the six months prior to the start of the treatment episode. Race/ethnicity was available only in Medicaid sample, so could not be included as a covariate, but was included in descriptive analyses.

### 2.4 Analyses

Analyses were conducted with SAS enterprise 7.1. Descriptive statistics were used to compare demographic information and prevalence of medication receipt between the two insurance cohorts. Because of low prevalence of medication receipt within the adolescent groups, insurance groups were combined for regression analyses and insurance status was treated as a covariate. Only a small number of adolescents received methadone, so methadone treatment episodes were not included in regression analyses. Further, because few adolescent treatment episodes were associated with extended-release naltrexone, we combined oral and extended-release naltrexone into a single category for regression analyses. Of note, approximately 75% of the naltrexone group was comprised of oral naltrexone. Thus, treatment types included in regression analyses were psychosocial services only, naltrexone (oral or extended-release), and buprenorphine.

We used logistic regression to estimate predicted probabilities of six-month retention by age group while adjusting for covariates. The predicted probabilities were equivalent to population margins based on covariates as observed in the sample and are also referred to as the adjusted prevalence. We first estimated six-month retention combining all treatment groups and accounting for covariates enumerated above with the exception of treatment type. We then stratified analyses by treatment type to estimate the odds ratios and adjusted prevalence for retention for each age group. Analyses were conducted using the SAS “surveylogistic” procedure to account for clustering of treatment episodes within individuals. The “lsmeans” option was used to obtain adjusted prevalence estimates.

To examine the effect of treatment type within each age group, we stratified analyses by age group and estimated six-month retention prevalence for each treatment type while adjusting for covariates.

We performed two sets of sensitivity analyses. First, to address the possibility that six-month continuous insurance required for primary analyses may affect results, we repeated analyses using three-month retention as the outcome variable. For these analyses, it was required that treatment episodes be associated with only three months of continuous medical and pharmacy coverage, starting from the first date of the treatment episode. Second, using co-morbid SUDs and co-morbid psychiatric disorders as markers of OUD complexity, we stratified sample based on median number of co-morbid SUD and psychiatric disorders and repeated analyses for six-month retention with a “high co-morbidity” and a “low co-morbidity” group.

## 3. Results

### 3.1 Sample

There were 629,480 individuals aged 12-64 with a diagnosis of OUD and OUD treatment claim in the database. After applying exclusion criteria, 277,084 treatment episodes were included in descriptive analyses.

### 3.2 Demographics

Table 1 shows demographic information for the sample. The most striking findings were age and insurance disparities in medication receipt. Approximately one-third (36%) of all Medicaid treatment episodes were associated with any medication, compared to 73% of commercial insurance treatment episodes (χ^2^ =57,870.6, *p*<.001). Furthermore, adolescents were less likely than adults receive OUD medication: within the commercial group, only 32% of adolescents received medication compared to 68% of young adults and 78% of older adults (χ^2^=5,027.8, *p*<.001); within the Medicaid group, fewer than 10% of adolescents received medication compared to 31% of young adults and 38% of older adults (χ^2^=921.8, *p*<.001).

Men composed the majority of commercial insurance group (62%); women composed the majority of the Medicaid insurance group (65%). Race/ethnicity information was available only for the Medicaid group where the majority of treatment episodes (83%) were associated with Caucasians. Co-morbid SUDs and other psychiatric conditions were most prevalent within the adolescent groups. Adolescents were less likely than adults to have an OUD diagnosis prior to the current treatment episode.

### 3.3 Regression Analyses

#### 3.3.1 Association between Age and Six-Month Retention

There were 261,356 treatment episodes included in regression analyses. Figure 1 shows the adjusted prevalence for six-month treatment retention by age group; the leftmost series shows the results for the entire cohort without adjusting for treatment type while the remaining series show results stratified by treatment group. Full results from the models are included in Supplementary Table 4. In the full cohort, there was a clear relationship between age and retention such that increasing age group was associated with increased retention: adjusted six-month retention prevalence for adolescents was 17.6% (95% CI 16.5-18.7%) compared to 25.1% (95% CI 24.7-25.4%) for young adults and 33.3% (95% CI 33.0-33.5%) for older adults. In contrast, after stratifying by treatment type, the association between age and retention was markedly decreased. For persons who received psychosocial services only, six-month retention was low for each age group (adolescents 13.9%, 95% CI 12.7-15.2%; young adults 12.9%, 95% CI 12.5-13.3%; older adults 15.8%, 95% CI 15.5-16.1%). Among persons who received naltrexone, adolescents were more likely to be retained in treatment compared to either adult group (adjusted prevalence for adolescents 32.7%, 95% CI 28.0-37.9%; young adults 18.4%, 95%

CI 17.3-19.5%; older adults 19.4%, 95% CI 18.5-20.2%). For persons who received buprenorphine, adolescents were equally likely to be retained in treatment compared to young adults but less likely to be retained compared to older adults (adjusted prevalence for adolescents 40.4%, 95% CI 36.9-44.0%; young adults 39.9%, 95% CI 39.4-40.4%; older adults 49.4%, 95% CI 49.1-49.8%).

#### 3.3.2 Association Between Treatment Type and Six-Month Retention

To formally compare retention across treatment groups within each age group, we estimated six-month retention prevalence for each treatment type stratified by age group (Table 2). Full model results are shown in Supplementary Table 5. Due to differing stratification variables—treatment type vs age group—prevalence estimates differed slightly between Figure 1 and Table 2.

Buprenorphine was superior to naltrexone and psychosocial services alone for each age group (Table 2). Naltrexone was superior to psychosocial services alone within each age group, although the degree of superiority decreased with increasing age: although naltrexone remained statistically superior to psychosocial services within the oldest age group, retention prevalence differed by fewer than 4% between the two groups.

#### 3.3.3 Sensitivity Analyses

##### 3.3.3.1 Three-Month Retention

Results for analyses estimating three-month treatment retention are shown in Supplementary Tables 6 (stratified by treatment type) and 7 (stratified by age group). An additional 43,392 treatment episodes with insurance coverage of at least 90 days but fewer than 180 days were included in these analyses. Overall, retention prevalence was higher at three months than at six months within each treatment group. We observed similar patterns for three-month retention as we did for six-month retention: in the combined treatment group, there was a significant age effect such that retention improved with increasing age; this effect was mitigated by medication receipt (Supplementary Table 6). Both naltrexone and buprenorphine were superior to psychosocial treatment within each age group, and buprenorphine was superior to naltrexone within each age group (Supplementary Table 7).

##### 3.3.3.2 Comorbidity Complexity

The median number of co-morbid SUDs and psychiatric diseases was one. Treatment episodes with one or zero co-morbid conditions (*n*=147,731) were therefore placed in the “low co-morbidity” group and treatment episodes with 2 or more co-morbid diseases (*n*=113,625) were placed in the “high co-morbidity” group. Results of regression analysis of six-month treatment retention are shown in Supplementary Tables 8 (stratified by treatment type) and 9 (stratified by age group). As with previous analyses, there was a significant positive association between age and retention in the combined treatment group which was markedly decreased when analyses were stratified by treatment type (Supplementary Table 8). There was a stronger association between medication and retention for the high co-morbidity group relative to the low co-morbidity group each type of treatment examined; this was true within each age group (Supplementary Table 9).

## 4. Discussion

In this large, retrospective study of OUD treatment episodes, we found clear age disparities in treatment and outcome: adolescents were significantly less likely than their adult counterparts to receive medication for OUD and were significantly less likely to be retained in treatment at six months. Importantly, we found these disparities were mitigated by medication treatment: when adolescents received medication, their retention rates improved and became similar or superior to those of adults. Importantly, while receipt of either naltrexone or buprenorphine was superior to psychosocial services alone within the adolescent population, buprenorphine was associated with significantly higher retention rates compared to naltrexone at six months. Buprenorphine was also associated with significantly higher treatment retention compared to naltrexone or psychosocial in each adult group studied.

Interestingly, naltrexone appeared to be more strongly associated with treatment retention within the adolescent group than within either adult group. One potential explanation may be that parental support during adolescence is helpful for ensuring adherence with daily medication. However, buprenorphine also commonly taken daily, yet buprenorphine’s association with retention did not decrease with increasing age. This may indicate that buprenorphine is more likely to be taken in absence of parental support than naltrexone, although future studies are needed to improve our understanding of the association between age and medication adherence.

Our results showing the high prevalence of psychiatric co-morbidities—both SUD and non-SUD— within the adolescent OUD population underscores previous findings that the diagnosis of OUD during adolescence indicates increased disease severity (Subramaniam et al., 2009; World Health Organization, 2009). Further, they support the assertion that this high-risk population should be offered the most effective treatment to mitigate the potential life-altering and life-threatening consequences of untreated OUD. Our findings suggest that pharmacologic treatment is superior to psychosocial treatment alone for treatment retention within the adolescent population, and provide further evidence that medication treatment should be offered to adolescents with OUD. This is of particular import given the low rates with which adolescents are prescribed medication, and given recent findings that the prevalence of buprenorphine prescribed to youth with OUD has decreased in recent years (Olfson et al., 2020). While the previously mentioned barriers to pharmacologic treatment for adolescents with OUD are numerous, our results add to the growing literature that points to need for increased access to medication treatment for adolescents with OUD (Committee On Substance Use and Prevention, 2016; Hadland, 2019).

Disturbingly, we found a significant insurance disparity in pharmacologic treatment for OUD. This was true in all age groups, but greatest in the adolescent population, where fewer than 10 percent of Medicaid treatment episodes were associated with any pharmacologic treatment. In contrast, 31% of adolescent commercial treatment episodes were associated with medication. Our data support recent findings buprenorphine is more commonly prescribed to those with commercial insurance compared to public insurance(Lagisetty et al., 2019) and further expands on them by demonstrating that this is also true within the adolescent population, and for all types of medication treatment. This finding is of considerable concern given the life-saving benefit of OUD medication (Larochelle et al., 2018; Morgan et al., 2019; Sordo et al., 2017) and points to the need for increased access to medication treatment for Medicaid-insured persons with OUD.

### 4.1 Limitations

The utilization of large administrative databases offers an important avenue to examine real-world outcomes in very large samples. However, due to the nature of administrative data, our variables were limited to what was available in the database. Although our sensitivity analyses largely confirmed our primary analyses, we cannot rule out that there were other unobserved variables or selection mechanisms that affected our results. For example, we were unable to adjust for race/ethnicity in our analyses, and there may be other important variables for which we could not adjust.

Due to the low prevalence of methadone receipt within the adolescent group, we were unable to perform regression analyses for the methadone treatment group. For similar reasons, we could not analyze persons who received extended-release naltrexone separately from those who received oral naltrexone. As the majority of persons within the naltrexone group received oral naltrexone, it is difficult to draw conclusions about the effectiveness of extended-release naltrexone from our results.

Importantly, the denominator of our study were persons with a treatment claim for OUD, and thus persons with undiagnosed OUD and persons with an OUD diagnosis but without an OUD treatment claim were not included. Thus, our results should not be interpreted as representative of the prevalence of medication receipt or treatment retention for OUD at the population level. Indeed, it has been estimated that fewer than 10% of all persons with OUD in the United States receive medication treatment (Williams et al., 2019), highlighting an important gap in implementation of evidence-based treatment in the OUD population.

## 5. Conclusions

Our results demonstrate that when given medication treatment, adolescents with OUD are similarly retained in treatment compared to their adult counterparts, and that buprenorphine in particular is associated with highest levels of retention. In addition, we found clear insurance disparities in medication receipt. Our results underscore the need for increased implementation of medication treatment for all persons with OUD, regardless of age or insurance status.

## Data Availability

Data are commercially available through Truven Health Analytics.

## Abbreviations

OUD: opioid use disorder
ICD: International Classification of Diseases
HCPCS: Healthcare Common Procedure Coding System
NDC: National Drug Codes
SUD: substance use disorder
CI: confidence interval

